# The D15 color arrangement test retains its diagnostic value regardless of display accuracy: A modeling study

**DOI:** 10.1101/2024.11.15.24314633

**Authors:** Tamás Árpádffy-Lovas, Edit Tóth-Molnár

**Affiliations:** Department of Ophthalmology, Albert Szent-Györgyi Medical and Pharmaceutical Center, University Szeged, Hungary, 6720 Szeged, Korányi fasor 10–11

**Keywords:** digital displays, color accuracy, color vision testing, color arrangement test, D15, deltaE

## Abstract

Color vision testing is often considered a resource-intensive examination, requiring an anomaloscope, or at least color plates and specialized lighting equipment. Multiple novel, digital color vision testing algorithms have also been described, but were only verified using calibrated displays. In this work, we assessed if color vision testing could be performed with any computer display using two arrangement type color vision tests, the D15 and desaturated D15 panels. Established mathematical models of human color vision deficiencies (CVDs) and publicly available colorimetric data of real world computer screens were used to model how the panel colors would be displayed on computer screens and then perceived and arranged by subjects with varying levels of CVD. A total of 627,431 arrangements were evaluated. We found that all of the modeled displays, including ones with a low color accuracy of deltaE=6, were characterized by specificity above 86%, and the sensitivity of the desaturated D15 panel remained above 85% as well, with negative predictive values above 90% to detect any form of CVD compared to using the unmodifed colors. These findings suggest that any digital screen could be utilized for color vision screening, even without calibration, enabling reliable high-throughput screening utilizing equipment that is already present at virtually all healthcare providers. Since 8% of the population is affected by some form of CVD, simplifeid screening would unlock notable benefits for all stakeholders. Furthermore, the described algorithm could also be used to optimize other digital color vision tests for general use.

## 1 Introduction

### 1.1 Color arrangement tests for color vision assessment

Color arrangement tests were first introduced in 1934 (Pierce). The modern and currently most utilized FM 100, D15, and desaturated D15 (D15-DS) panels were originally described by Farnsworth and Munsell (1943), and Lanthony (1974), respectively.^1^

These tests are used to evaluate color vision, and can be utilized for screening purposes as well as a part of the ophthalmological diagnostic toolkit. High-throughput screening for color vision deficiency (CVD) is needed in various populations, for example, the D15 panel is currently in use for screening purposes at the Canadian Armed Forces.^2^

The technical requirements of color arrangement tests include a standard light source, preferably a D65-equivalent lamp, and the color caps, which need to be handled carefully to avoid degradation. When the test is performed, the caps are manually ordered, then manually randomized, and then ordered again by the subject by choosing the color that is the most similar to the previous one, starting from the fixed first (“P”) cap. This is followed by turning over the caps to write down their order by the examiner. Since color arrangement tests need to be taken at least twice to optimize their diagnostic precision,^3,4^ these steps are repeated multiple times for each patient, requiring at least 10 minutes if each eye is tested individually. Non-standard environments may also negatively affect the performance of patients in color arrangement tests.^5^

### 1.2 Scoring algorithms

The D15 and D15-DS panels consist of 16 colored caps. The zeroth cap, the P cap, is fixed and cannot be moved. The numbered caps range from 1 to 15, and these caps are arranged by the subject by placing the cap with the most similar color to the previous one.

The arrangements can be evaluated with a qualitative, visual approach. In this method, the CVD type is identified by predefined axes of confusion, and counting the number of color swaps made by the subject are used to estimate severity. These qualitative approaches are less time consuming compared to more complex calculations, but their reliability might be limited due to the introduction of subjective elements to the diagnosis.^8^

The calculations in this work are based on two quantitative approaches, the moment inertia method described by Vyngris and King-Smith^9^ (referred to as VKS; 1988) and the least squares linear regression technique described by Foutch, Stringham, and Lakshminarayanan^10^ (referred to as FSL; 2011).

Both of these methods utilize the same canonical colors for the D15 and the D15-DS caps with the D65 standard illuminant, defined in the 1976 CIELUV colorspace. As the brightness of the illuminant can be considered the same for each cap, the L value is not used in the equations. The u’ and v’ values represent the hue of each color on a red-green and a blue-yellow axis, respectively.

If the panel colors are not represented correctly on a digital screen, this will result in discrepancies in the u’ and v’ values of the colors, but only in the representation; when the color arrangement is evaluated, still only the canonical colors can be used by the scoring algorithm in order to obtain comparable and repeatable results.

This leads to a notable ambiguity in the evaluation: without display calibration, one can never be sure if the colors seen and arranged by the patient were the same as the ones used for scoring the arrangement. However, even a perceivable difference in color might be acceptable.^7^ Most notably, if the color difference does not lead to color swaps, i.e., the change in colors does not affect the expected arrangement of the color set. Therefore, the amount of acceptable difference in color may be quantified.

### 1.3 Color difference (deltaE)

Difference in colors or color representations is expressed in deltaE (dE) values. Multiple algorithms in different colorspaces have been described for its calculation.^11^ Digital displays are most often characterized by their dE2000 value, based on the CIELAB colorspace.^12^ According to a whitepaper on color management by the International Color Consortium (ICC), when comparing two colors for at least 15 seconds, a difference of dE=5 can be identified by untrained eyes.^13^ It has also been suggested that dE<2.5 may not be detected and dE<1 cannot be detected by untrained observers. Here we consider dE<1.0 to be the level of undetectable color difference.^13^

### 1.4 Digital displays in color vision assessment

With the advent of accurate digital displays, the digital representation of the same colors have been assessed as a possible replacement of the physical tests. The main concern regarding digital color vision testing is the uncertainty of the diagnostic value of the test, if the color accuracy of the display used to take the test does not match the required level.^6,7^ The first color vision test type to be utilized digitally were variations of pseudoisochromatic image tests, including the Cambridge Color Test, the Rabin Cone Contrast Test, the Richmond HRR, and the Konan– Waggoner Computerized Color Vision Test.^7,14^ A new digital and automated color vision test has also been recently described by Fanlo-Zarazaga et al., also based on pseudoisochromatic images.^15^ An advantage of digital testing is that, unlike the commonly utilized Ishihara plates, pseudoisochromatic images generated randomly on-the-fly cannot be memorized before taking the test. Multiple smartphone-based studies also compared digital pseudoisochromatic images to printed Isihara plates. Some authors have reported encouraging levels of agreement between the two methods,^16,17^ while others have reported much lower diagnostic value of smartphone applications compared to the printed plates.^18^ All of the above works have concluded that both smarthpone applications and smartphone displays would require improvements to become clinically viable alternatives of standard Ishihara plates. Even though a thorough colorimetric analysis of tablet and cell phone device screens has shown that their average color accuracy cannot be considered high enough for color critical activities,^19^ in this work we intend to demonstrate that such precision may not be required for arrangement type color vision testing.

Almustanyir et al. have also compared digital pseudoisochromatic tests with digital and standard versions of the D15 panel and have found that the Konan–Waggoner Computerized D15 panel (KWC-D15) could substitute the standard D15 panel.^14^ Furthermore, they have also demonstrated that the KWC-D15 color vision test offers high clinical diagnostic value on two specific devices.^20,21^

However, thorough clinical evaluation of numerous digital displays would require a substantial amount of time as well as a relatively high number of subjects with CVD diagnoses established using the still gold standard Nagel anomaloscope. Instead, here we focus on a modeling approach that provides an approximation of such clinical testing.

## 2 Theory and calculation

### 2.1 Programming approach

The project consists of custom Python scripts, depending on the Colorspacious, Colormath, NumPy, SciPy, and Matplotlib libraries. The scoring algorithms were also implemented in Python as described in their respective publications.^9,10^ The whole project is available on GitLab for others to use and modify under the GPLv3 license.^23^

### 2.2 Simulated CVD

Machado et al. defined a physiologically-based algorithm to simulate the color appearance of anomalous trichromat observers using the spectral response of cone pigments that can be utilized to simulate CVD in images.^24^ A similar algorithm has also been reported to be applicable for predicting the D15 arrangement of real-life observers with CVDs.^25^ The implementation of the algorithm in the Colorspacious Python library was used to alter the D15 and D15-DS panel colors as if they were perceived by observers with protanomaly, deuteranomaly, or tritanomaly of 0–100% (Figure 1, Figures 2C and 2G). The percentages represent the loss of functionality of each cone and thus the expected perception of each color. The severity of the CVDs was set in 1% increments, therefore a total of 303 observers were modeled in this manner.

**Figure 1.**
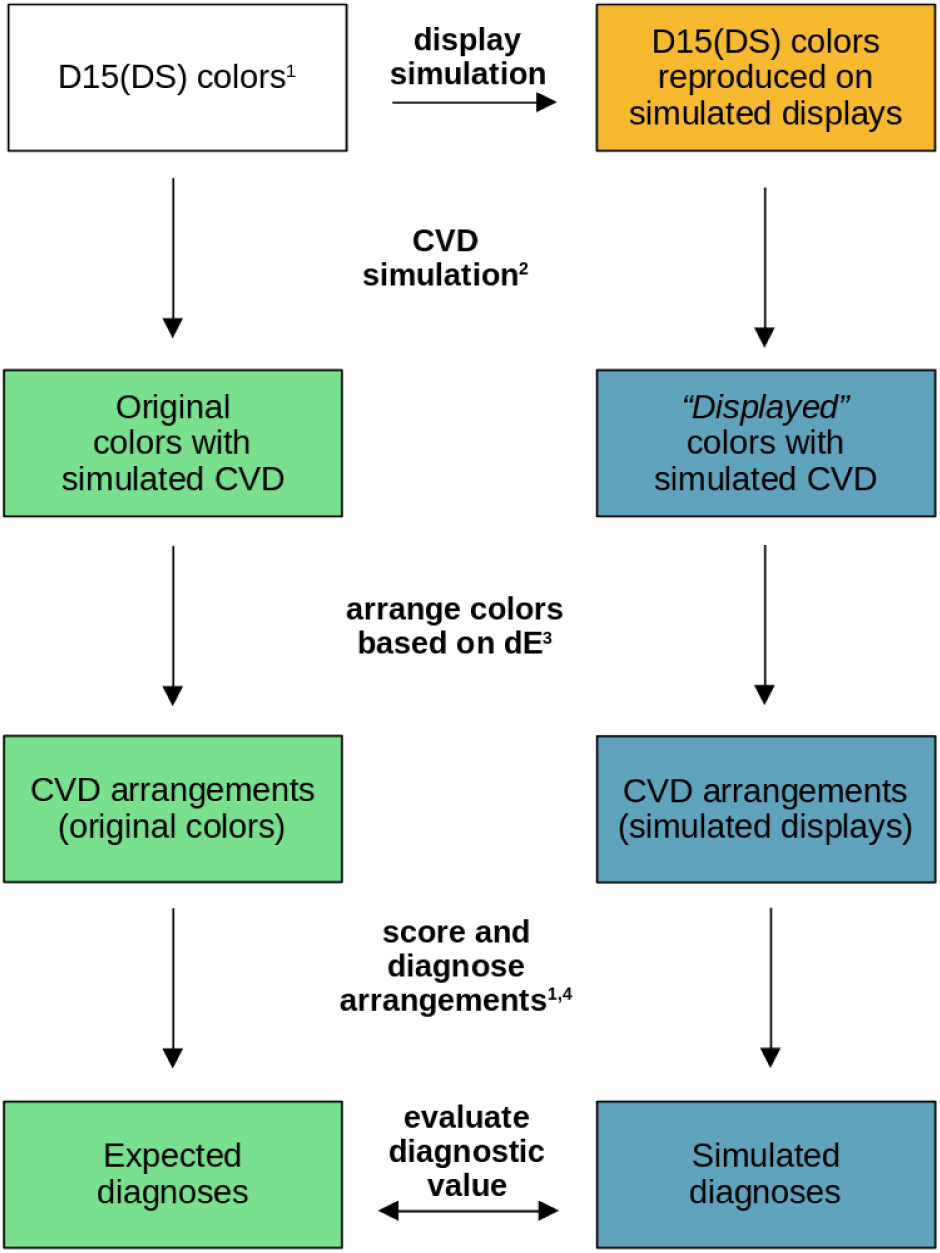
Steps of the simulation. The numbers refer to the following articles: 1, Vyngris and King-Smith, 1988; 2, Machado et al., 2009, 3, Almustanyir et al., 2021; 4, Foutch et al., 2011

**Figure 2.**
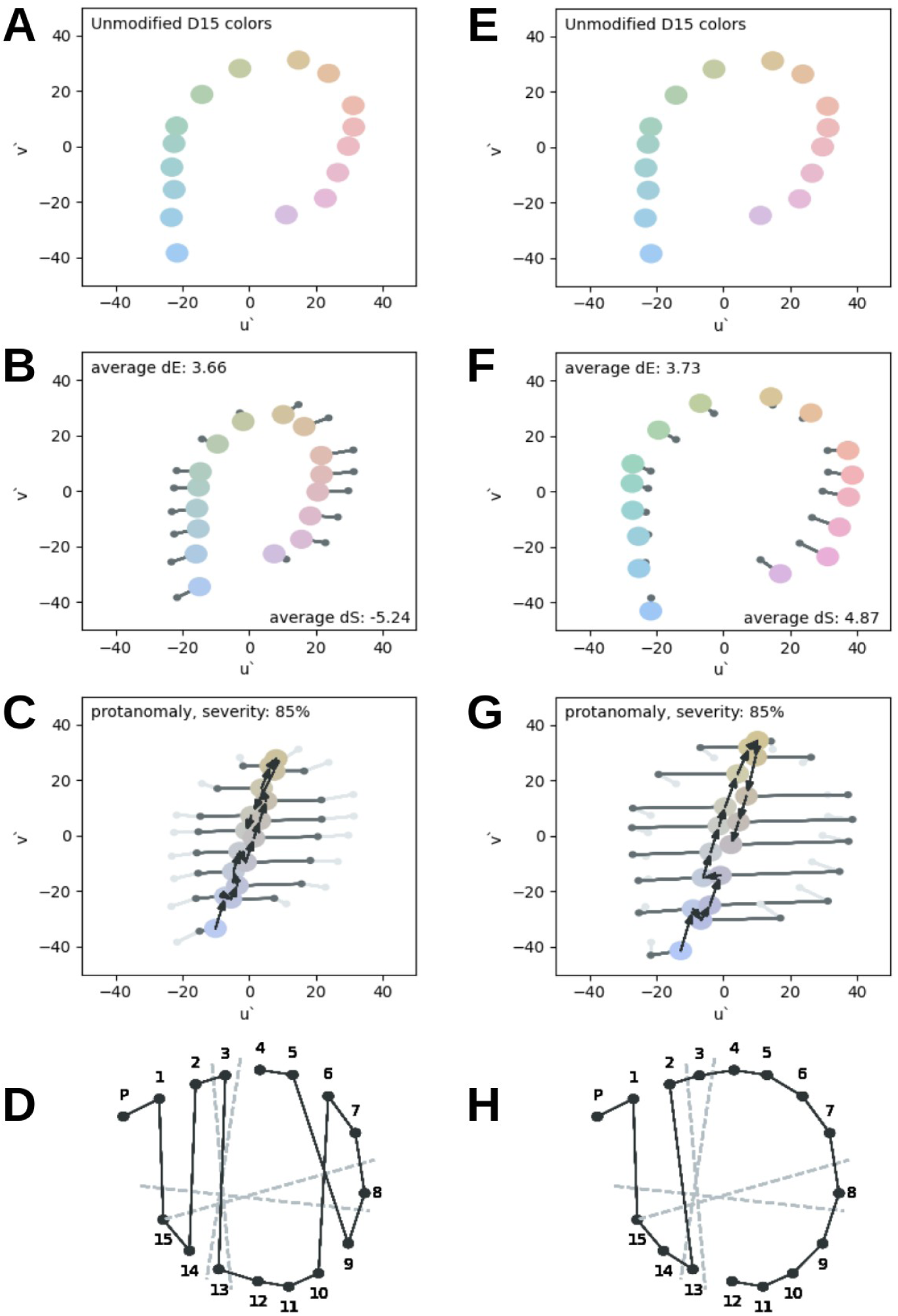
Representative positions of the normal D15 colors (A and E), and the changes in their positions after applying display profile modeling (B and F) and color vision deficiency modeling (C and G) in a desaturating (left) and an oversaturating (right) display, where black arrows show the steps of arranging the colors based on for others to use and modifythe lowest dE values. D and H are the qualitative representations of the arrangements corresponding to C and G, respectively.

The modeled observers performed color arrangement by selecting the color that had the lowest dE value compared to the current one, starting from the fixed P cap, similarly to the algorithm described by Hovis and Almustanyir.^25^ The CAM02-UCS uniform colorspace dE calculation from Colorspacious was used to find this lowest dE value between subsequent colors.^26^

Original diagnoses are the diagnoses obtained by performing the D15 and D15-DS arrangement tests, but only using the unmodified colors that were originally reported by Vyngris and King-Smith^9^ with modeling the corresponding CVD, but without modeling display color reproduction. The resulting arrangements were evaluated with both the VKS and FSL scoring algorithms (Figure 1, left side of the chart).

### 2.3 Display color reproduction model

Display color accuracy can be evaluated by displaying a set of standardized calibration color values and measuring the color output of the display using a colorimeter. Then, all content for the given display can be optimized with this profile by matching the color gamut of the content to the colors that can be reproduced by the display. Equation 1 shows the mathematical approach for matching a given color in the sRGB colorspace to the XYZ values optimized for the display.

Colorimetric profiles and measured average dE values were obtained for the displays of notebook computers released between 2022 and 2024 from NotebookCheck.net^27^ in the ICC file format.^28^ These measurements have originally been performed for review purposes and the resulting ICC profiles can also be used to improve the color accuracy of the corresponding laptop computer with the same display.

To model the color representation of each display, a total of 52 target colors were utilized. Two color sets contained the D15 and the D15-DS panels, defined in the 1976 CIELUV colorspace ^12^ and converted to the sRGB colorspace with an L value of 80 and D65 standard illuminant whitepoint. The third color set contained the Color–Rendition Chart^29^ colors in the sRGB colorspace.

The intended use for Equation 1 is a one-way transition of the red, green, and blue subpixel intensities of a selected color to the value of the same color optimized for the profile of the given display.^13^

To model the expected representation of a color on the uncalibrated screen, we need to find the red, green, and blue input values for *Lr*, *Lg*, *Lb* that will result in the target color. This optimization was done using the SciPy implementation of the Nelder–Mead algorithm.^30^

In this manner, an expected reproduction of the Color–Rendition Chart^29^calibration color set and an expected reproduction of the D15 and D15-DS panel colors were generated (Figure 1 right; Figure 2, second row). The average dE2000 value of the profile-fitted calibration colors was used to compare our modeled color accuracy to the one measured using a colorimeter.

The algorithm described by Machado et al.^24^ was then applied to the profile-fitted D15 and D15-DS panel colors to model their perception by CVD observers (Figure 2, third row). These color values were then arranged starting from the P-cap by choosing the closest color based on dE value (Figure 2, fourth row). In this case, the CAM02-UCS colorspace dE was used instead of dE2000. Furthermore, if two colors were closer than dE=1 to each other, their difference in color was considered to be imperceptible, and such colors were used to generate additional arrangements that consisted of swapping all of the perceptually similar colors, further increasing the total number of evaluated arrangements.

Finally, the diagnosis of these color arrangements were compared to the original diagnoses of the corresponding simulated observers, enabling the evaluation of diagnostic value of a test performed on each display.

### 2.4 Difference in saturation

Similarly to the dE value, the difference in saturation between two colors was also defined. The deltaS (dS) value is defined as the difference in chroma values in the JCh colorspace, also part of the CIECAM02 model.^26^ Therefore, the distance of the two colors in dS is a lot less affected by hue or brightness. This value is used to quantify the possible oversaturating or desaturating effects of displays (Figure 2, B vs F and C vs G).

### 2.5 Statistical analysis

Statistical analysis was performed with custom scripts, figures were created using Matplotlib. The measured and estimated dE2000 values for the profile-fitted colors were compared in a Bland–Altman plot. Spearman correlation and linear regression fitting were performed where applicable, p<0.05 was considered statistically significant.

The diagnoses of the simulated color arrangements were compared to the original diagnoses for each modeled observer, and marked as true positive, true negative, false positive, or false negative. These evaluations were performed both specifically for each anomaly as well as in a general manner. The general classification considered the detection of any anomaly a true positive, regardless of anomaly subtype. Specificity, sensitivity, negative predictive value (NPV), and positive predictive value (PPV) were calculated as defined by Shreffler and Huecker.^31^ Then, the diagnostic value of each modeled display was described by its sensitivity, specificity, accuracy, positive and negative predictive value; these metrics were considered acceptable for use in a clinical setting when their values were >=90%.

## 3 Results

### 3.1 Modeling color reproduction

A total of 452 ICC profiles were acquired, 247 of which were accepted as an adequate model for further investigation. A display was considered adequately modeled when the average fit precision was dE<1.0, and the average dE value of the calibration color set was within ±50% of the originally measured dE values of the corresponding display reported by Notebookcheck.net.^27^

The changes in the positions of individual colors in both calibration and D15/D15-DS color sets were plotted in the CIELUV 1976 colorspace for each screen. For brevity, these three plots for each individual display are not included here, but can be reproduced by running the script. Figure 2 shows the steps of the calculations from the original colors through the colors on the simulated display to the colors perceived by a modeled observer with 85% protanomaly.

In general, the most common observed change in the modeled color reproduction, compared to the original colors, was either an overall decrease or an overall increase in color saturation, while hue directions from the origo were similar. In Figure 3 left, a Bland–Altman plot shows that, on average, the dE value of the displays was 1 lower when profile-fitting the colors using the method explained in 2.3, and the difference increased as the measured dE values of the displays increased. On the other hand, the difference in the average dE values calculated from the modeled reproduction of the the D15/D15-DS panel color sets was, on average, dE<0.5, and the overall spread of the differences was dE<1.

**Figure 3.**
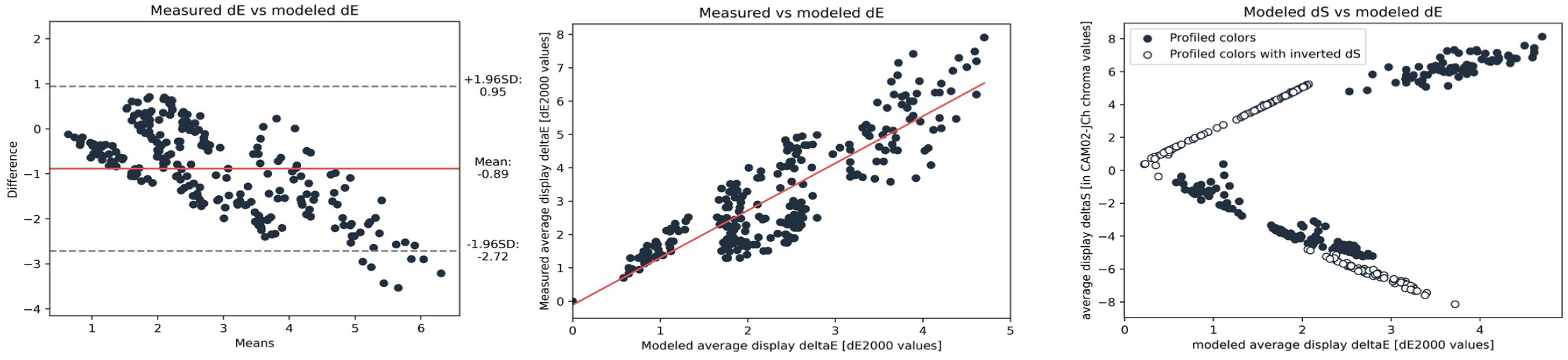
Left: Bland–Altman plot showing that dE calculations based on the calibration color set generally underestimated the measured dE; middle: Scatterplot showing the linear association (red) between measured and modeled dE values; right: Scatterplot showing the interaction between dS and dE values in modeled displays (dark) and the dE values of the modeled displays with inverted dS (light).

If we wanted to predict the measured dE value of each display based on the modeled reproduction of the calibration color set, the modeled average dE value would be falsely low in most displays (Figure 3 left and middle). A possible explanation of this phenomenon is that our calculations were performed on a limited number of standard calibration colors, while the colorimetric measurements also included numerous colors, including ones at the edges of the target display gamut.

Nevertheless, as Figure 3 middle shows, a linear association was present between the calculated and the modeled average dE values (p<<0.01); therefore, to maintain the association shown in Figure 3 and still provide predictions on real-world display performance at the same time, linear regression was used to match the modeled and the measured dE values. The diagnostic values (Figures 4 and 5, Supplementary Material) are to be shown with these estimated dE values for each display.

**Figure 4.**
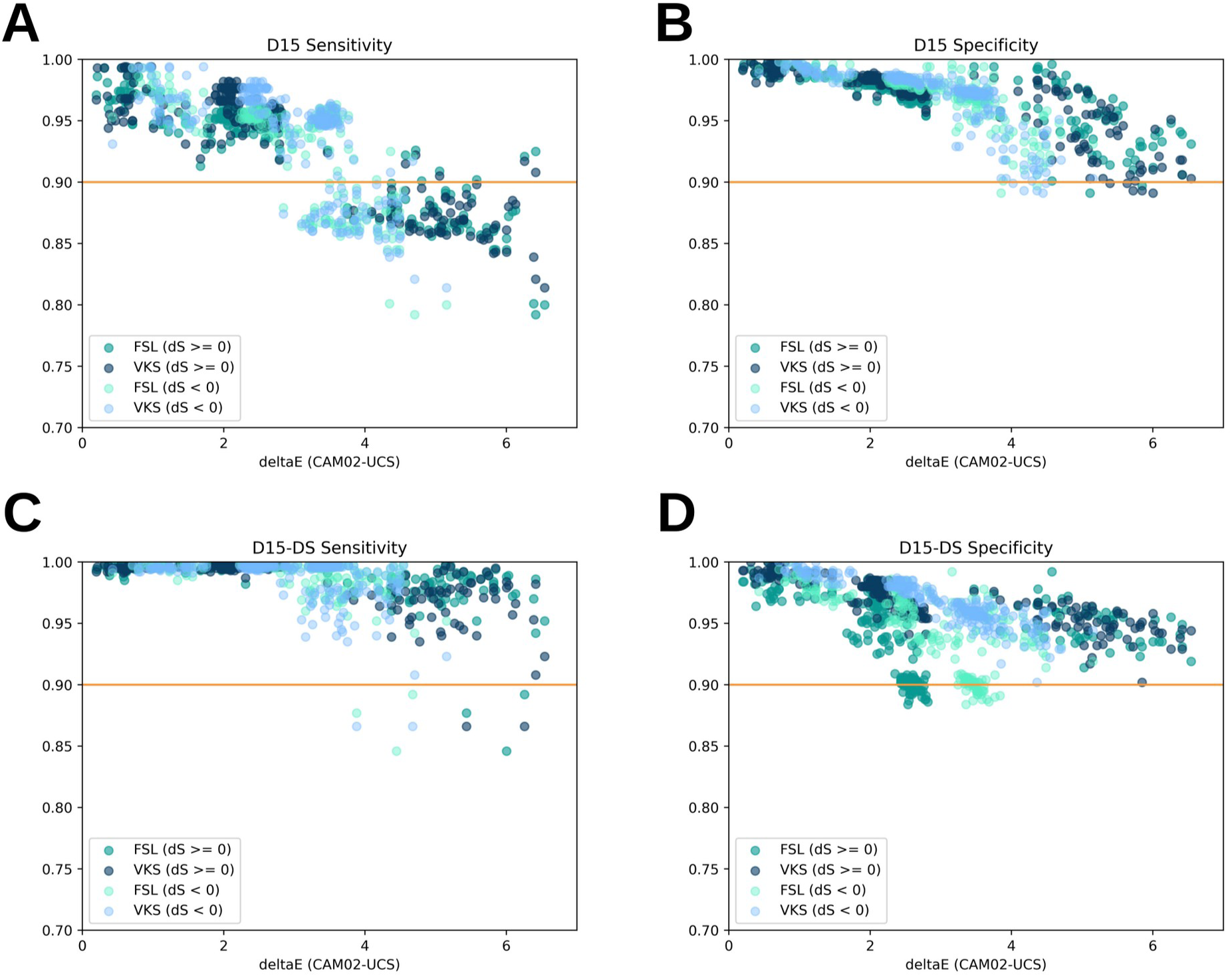
Scatter plots showing that both the sensitivity (A, C) and the specificity (B, D) of both D15 (A, B) and D15-DS (C, D) panels decreased with the increase of dE, but the decrease was more pronounced in displays with dS<0. Orange lines represent the limit of 0.9.

**Figure 5.**
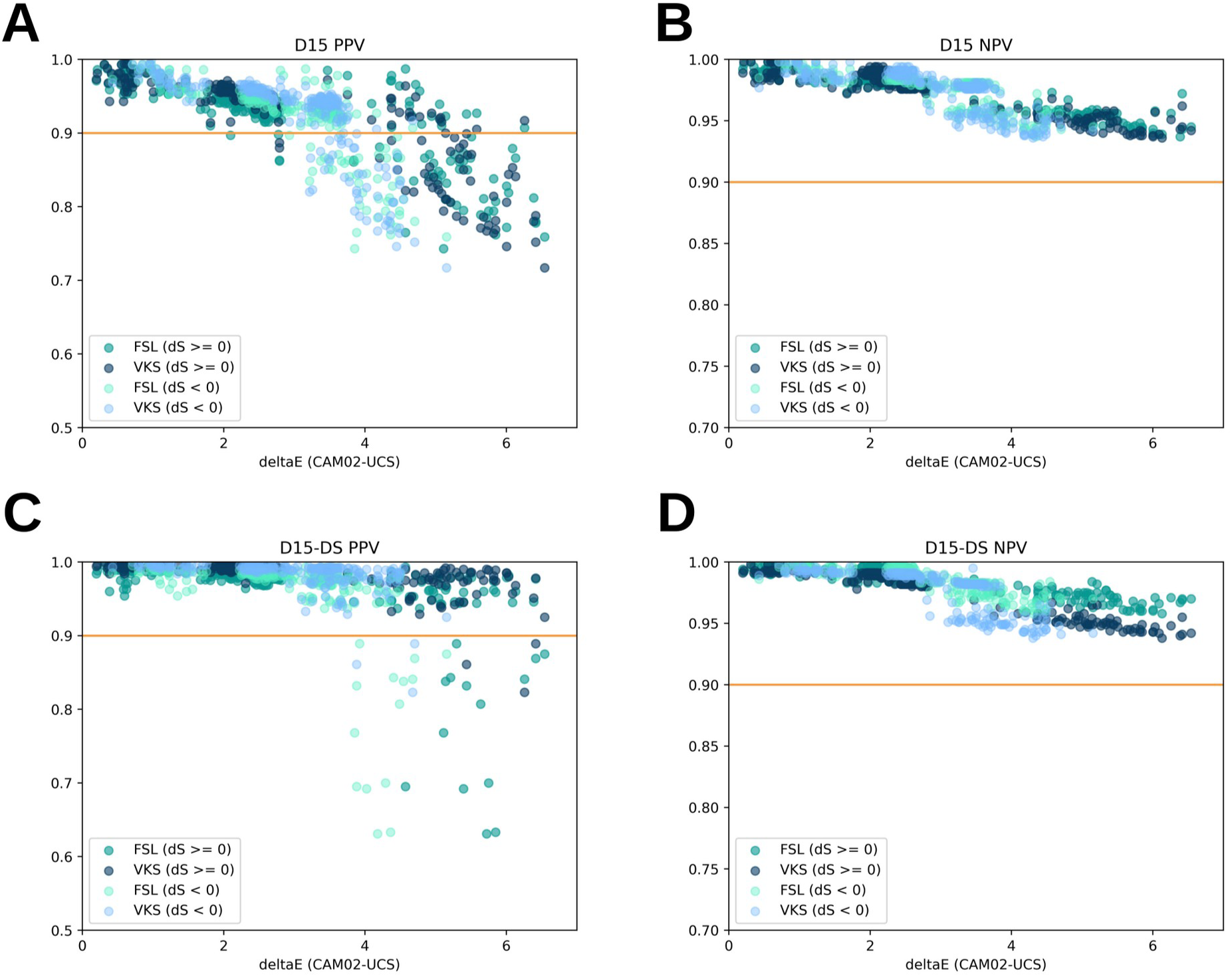
Scatter plots showing that both the positive predictive values (A, C) of both D15 (A) and D15-DS (C) panels decreased with the increase of dE, but the decrease was more pronounced in displays with dS<0, while the negative predictive value was only slightly affected by the increase of dE in the D15 (B) and the D15-DS panels (D). Orange lines represent the limit of 0.9.

### 3.2 Changes in color saturation

As mentioned in 3.1, the display display simulations caused either an overall increase or an overall decrease in saturation, expressed as dS. In the evaluated display profiles, the saturation decreased in displays with dE<2, and increased mostly in displays with dE>2. However, the dE and the dS metrics do not need to be associated in such a way. Therefore, all of the display simulations were repeated once more, and in this additional step the change in saturation from the target color to the profile fitted color were inverted: the extent of change in dS was maintained, but the direction of the change was reversed. This removed the bias towards negative dS being associated with lower dE values, and also doubled the number of simulated displays (Figure 3 right).

### 3.3 Simulated CVD observers at baseline

The D15 and D15-DS colors were arranged starting from the “P” cap after CVD simulation, based on their relative dE values (Figure 1, left column; Figures 2C, and 2G), and these arrangements were assigned a diagnosis based on their scoring with the VKS and FSL algorithms. Table 1 shows the cutoff values in severity where each simulated CVD was detected.

**Table 1.**
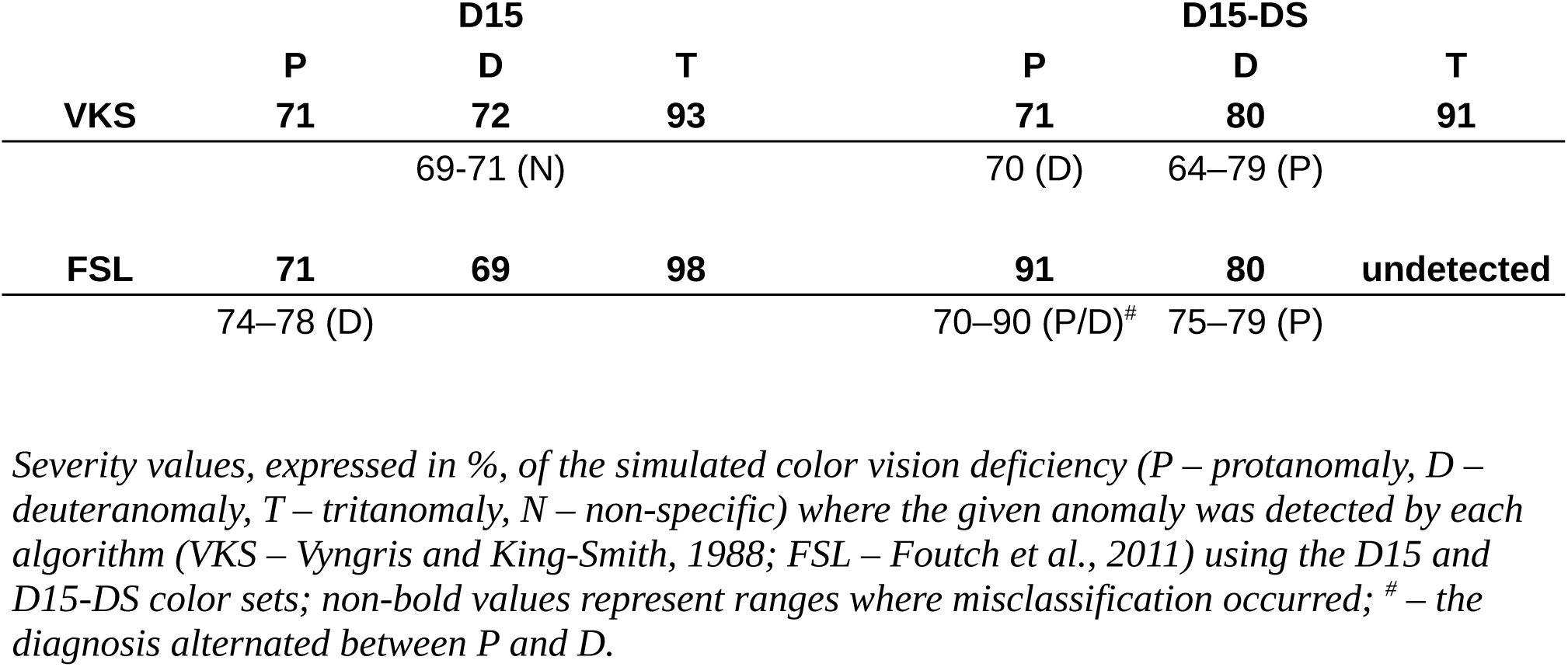

Simulated protanomaly was detected from 71% severity with the D15 color set by both algorithms, although FSL reported deuteranomaly in the range of 74–78%. With the D15-DS colors, VKS reported deuteranomaly only at 70%, and the diagnosis was correct from 71%. On the other hand, the FSL algorithm stably reported protanomaly only from 91%, while returning protanomaly or deuteranomaly interchangeably from 70%.

Deuteranomaly was also correctly detected with the D15 colors by both algorithms, only VKS returned non-specific anomaly between 69–72%. Conversely, with the D15-DS colors, simulated deuteranomaly was misclassified as protanomaly from 64% (VKS) and 75% (FSL) up to 80%, from where it was correctly detected.

While protanomaly and deuteranomaly were detected in the 71–80% range, simulated tritanomaly remained undetected below 91% severity, where it was detected by the VKS algorithm with the D15-DS colors, and at 93% with the D15 colors.

Simulated tritanomaly was completely undetectable by the FSL algorithm with its originally reported parameters. The authors recommend that confusion angle (θ) values above 70° should be diagnosed as tritanomaly, but this angle was too steep to detect the simulated tritanomaly. In addition, arrangements that with qualitative analysis showed definite tritanomaly of some level were also diagnosed as unspecified CVD due to the corresponding angle being below 70°. Therefore, in this work θ>49° was considered as tritanomaly. Despite this change enabling the VKS algorithm to detect tritanomaly at 98% severity with the D15 colors, tritanomaly remained undetectable with the D15-DS colors.

When the profile-fitted D15 and D15-DS colors were evaluated, the expected diagnosis was the exact diagnosis corresponding to the given algorithm, color set, anomaly, and severity. This also includes misclassifications: if the diagnosis was the same, it was considered a true positive even if the original diagnosis classified the simulated anomaly incorrectly. Therefore, we are focusing on receiving the same diagnosis with and without display modeling, and not diagnosis correctness on its own.

### 3.4 The effect of simulated display color accuracy on overall diagnostic value

A total of 627,431 individual arrangements were evaluated. The following results represent the detection rate of any anomaly where anomaly was present, disregarding any misclassifications of anomaly subtypes. The Supplementary Material contains the diagnostic accuracy values for all three CVD types individually.

The general sensitivity of both algorithms with the D15 colors remained above 90% up to dE=2, and showed a decrease to 80% above it (Figures 4A and 4C). Even though both the increase and the decrease in saturation resulted in similar sensitivity in the dE>2 range, sensitivity markedly decreased at higher dE values in the displays that were characterized by decreased saturation (dS<0, Figure 4A). Other displays that were characterized by dS>0, sensitivity remained above 0.9 up to dE=3.5 with the D15 colors. On the other hand, sensitivity with the D15-DS colors was below 0.9 only in 10 screens at and above dE=4 (Figure 4C).

The D15 colors resulted in specificity values above 0.85 with all screens and both algorithms regardless of dS values (Figure 4B). The specificity with the D15-DS colors remained above 0.9 up to dE=2, and showed a a slight decrease but remained above 0.85 from there all the way to dE=0.6 (Figure 4D).

The overall PPV remained above 0.9 up to dE=2 and up to dE∼4 with the D15 and the D15-DS colors, respectively (Figures 5A and 5C). Decreased saturation negatively affected the PPV of both color sets, as screens with dS>0 performed markedly better in this regard. The PPV of the D15-DS set remained generally more accurate.

Both algorithms and both color sets produced NPV values above 0.9 in all simulated screens, with the FSL algorithm being slightly superior with the D15-DS colors in this regard (Figures 5C and 5D). Simulated displays that increased saturation produced higher NPVs, but the general performance of this metric can be considered unaffected by the direction of saturation change.

While overall clinical diagnostic values remained relatively high in the detection of any CVD up to the dE=4 point, especially with the D15-DS color set, correct classification of anomaly type was less reliable. The D15 color set was less affected by dE in specificity, as it remained above 88% up to dE=6, but sensitivity was markedly decreased to around 50% from dE=4 in protanomaly. The D15-DS colors had similar performance in sensitivity both in deuteranomaly and protanomaly, but specificity was decreased sharply all the way to 20% at dE=4. Specificity in modeled observers with tritanomaly remained above 90% at all dE values with dE colors, but sensitivity was poor, even in very color accurate displays. Overall, correct discrimination between protanomaly and deuteranomaly allowed for only moderate changes in saturation, and, subsequently, dE values below dE∼4. Tritanomaly was found to be difficult to detect using these color arrangement tests even without display modeling.

## 4 Discussion

We evaluated the effect of display color accuracy on the diagnostic value of D15 and D15-DS color arrangement tests scored with the VKS and FSL algorithms and simulated on display profiles generated with colorimetry by simulating discreet severities of CVDs, and by simulating the color arrangements of CVD observers.

A number of studies have already shown the adequate performance of color arrangement tests using calibrated digital displays in a controlled clinical environment,^6,7,15,21,22,32^ and our results suggest that the D15 and the D15-DS color arrangement tests performed on digital displays are likely to be characterized by high sensitivity, acceptable specificity, high PPV, and exceptional NPV, making them ideal for screening purposes at any level of display color accuracy. Differentiation between protanomaly and deuteranomaly would still require specified equipment, but such specific diagnosis may not be required in all screening scenarios.

It should be noted that the high sensitivity and specificity correspond to a high level of agreement between the non-modified and the profiled color sets when modeling the color arrangements, therefore, this relates to the performance of the displayed colors compared to the original D15/D15-DS panels. While further studies with are still needed to understand the diagnostic performance of a generalized digital D15 test, real-world diagnostic performance would most likely be in the same ranges as previously reported: 0.32–0.58 and 0.51–0.79 for sensitivity, and 0.83–1.0 and 0.77–1.0 for specificity with the D15 and the D15-DS panels, respectively.^33–35^

The lowest level of detectable CVD with the simulated color arrangement was above 70% decrease in the function of the red and green cones, despite the fact that even a decrease of 50% would also lead to substantial change in color perception. This could be explained by the fact that human color perception is characterized by dynamic understanding of color differences, affected by other colors perceived in the environment, while the dE values calculated to generate the simulated arrangements are exact distances, unaffected by external factors in the context of the modeling approach.^36,37^ The psychophysical nature of color perception should be accounted for when designing digital versions of color vision tests by optimizing variables that may affect relative color appearance, such as background color and target display brightness. Despite this limitation, the consistency of the results above the threshold of detectable CVD indicates that the digital representation of the D15 and D15-DS colors can be viable substitutes of the original color caps, enabling quicker diagnostic procedures by automating the manual steps of setting up and evaluating the test.

It should be noted that even though the limit of dE<4.0 may be considered as a reasonable cutoff for choosing displays to be used for color vision testing, the change in saturation, dS, was a more pronounced factor in our model. Every modeled display was evaluated twice, once with the original direction of the induced saturation change, and once with the direction of the change in saturation inverted. Even though some displays in the 2–4 dE range were originally desaturating ones and others had the desaturating effect computationally introduced, all of the modeled displays that were characterized by desaturation performed worse in all metrics except the specificity with the D15 panel. On the other hand, the specificity and the PPV of the D15-DS panel was most affected by desaturation, but a lower number of modeled displays produced this effect. Tritanomaly, which is much less common compared to protanomaly and deuteranomaly, was correctly detected in very low rates even without any modification of the D15/D15-DS panel colors, possibly indicating that the more time consuming but also more precise FM-100 panel could be a more appropriate color arrangement test if tritanomaly is suspected.

The dynamic generation of random pseudoisochromatic images and random starting color arrangements might also aid to mitigate the limitations introduced by the displays used for color vision testing. Current advances in artificial intelligence and machine learning technology are already facilitating a possible digital transition in color vision testing with novel diagnostic approaches.^32^

Due to the fact that both the VKS and the FSL algorithms rely on the canonical colors for evaluation, dynamically choosing colors to arrange seems less feasible, unlike in pseudoisochromatic image testing. However, the colors used by both the D15 and the D15-DS panels have been defined as physically available caps more than 80 and 50 years ago, respectively. While digital alternatives have been available for some time, both in free and in commercial forms, these methods are either based on the original colors or utilize proprietary color choices. As the results in this work and the results of others^14,15^ show, both approaches may be viable for color vision testing on digital screens, but their general performance is difficult to evaluate. It would be beneficial to define a standard color set for color arrangement tests that could be openly implemented.

Dain and Adams have suggested that the colors of the D15-DS panel may not be optimized in terms of the dichromatic confusion loci, possibly decreasing the reliability of correct identification of a CVD type,^38^ which was also supported by the findings in the paper from Atchinson et al., where higher probabilty of misdiagnosis was noted for the desaturated panel.^22^ Due to the slight differences in diagnostic performance between the D15 and the D15-DS panels, it would be advisable to test subjects with both panels with a few minutes of rest between the to tasks, as this could decrease the influence of the diagnostic uncertainty introduced by possibly unknown display color accuracy. In this sense, applicable displays could include laptop and desktop computers, tablets, or even smartphones.

The pipeline described in this work could also be used to evaluate the diagnostic value of other original, digital-first color vision tests, which have been primarily relying on calibrated, very color accurate screens.^15,32^ With this approach, a display-agnostic test battery could be implemented that could include both pseudoisochromatic images and color arrangement tasks, possibly enabling a quick, even gamified approach to color vision screening and testing.

## 5 Conclusion

Our results suggest that, in light of the very high NPV achieved, virtually any digital display could be used for color vision screening with the D15 and D15-DS arrangement type color vision tests in a regular office setting, without any specified equipment. Furthermore, displays with dE<2 may even be accurate enough to determine CVD subtype and severity with reasonable precision. Obtaining such displays is enabled by numerous display manufacturers disclosing precise color accuracy metrics of displays in their marketing materials. In addition, the described algorithm could also advance the development of completely digital color vision testing methods.

## Data Availability

All data can be reproduced using the code uploaded to the project repository.

https://gitlab.com/ThomasHastings/d15-display-deltae

## 6 Acknowledgements

The authors would like to express their gratitude for the colorimetric data made publicly available by Notebookcheck.net, as it was integral to the calculations presented in this work. This work was funded by the University Research Scholarship Program of the Ministry for Innovation and Technology through the University of Szeged (EKÖP-24-4-367).

## 7 Author contributions

TÁ-L contributed to conceptualization and methodology, software development, data curation, formal analysis, validation, visualization, writing – original draft; ET-M contributed to funding acquisition, supervision, writing – review and editing.

## Equations

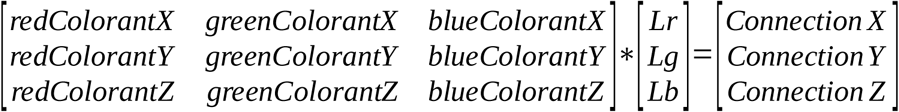

**Equation 1** Where Lr, Lg, and Lb are the red, green, and blue color components corrected for the corresponding gamma values of the display, and the 3×3 matrix contains the response curves of the display for each base color. These values can be obtained from the calibration data, i.e., the colorimetric profile, for any display.

## Notes

### Competing Interest Statement

The authors have declared no competing interest.

