## Supplementary_material for "The D15 color arrangement test retains its diagnostic value regardless of display accuracy: A modeling study"

This document contains detailed diagnostic parameters, including accuracy, sensitivity, specificity, as well as negative and positive predictive values (NPV and PPV) for the D15 and D15-DS color arrangement tests on simulated protan, deutan, and tritan observers with a total of 494 simulated digital displays.

The figures on the top left depicting the variables for all color vision defects combined are the same ones presented in the main article, except for accuracy, which is only shown here.

Abbreviations in the figures:
**FSL**, scoring algorithm described by Foutch et al. (2011)
**VKS**, scoring algorithm described by Vyingris and King-Smith (1988)
**deltaE**, difference in color, lower values correspond to a more color-accurate display
**dS,** change in saturation; if dS>0, the modeled display increased, and if dS<0, the modeled display decreased saturation.

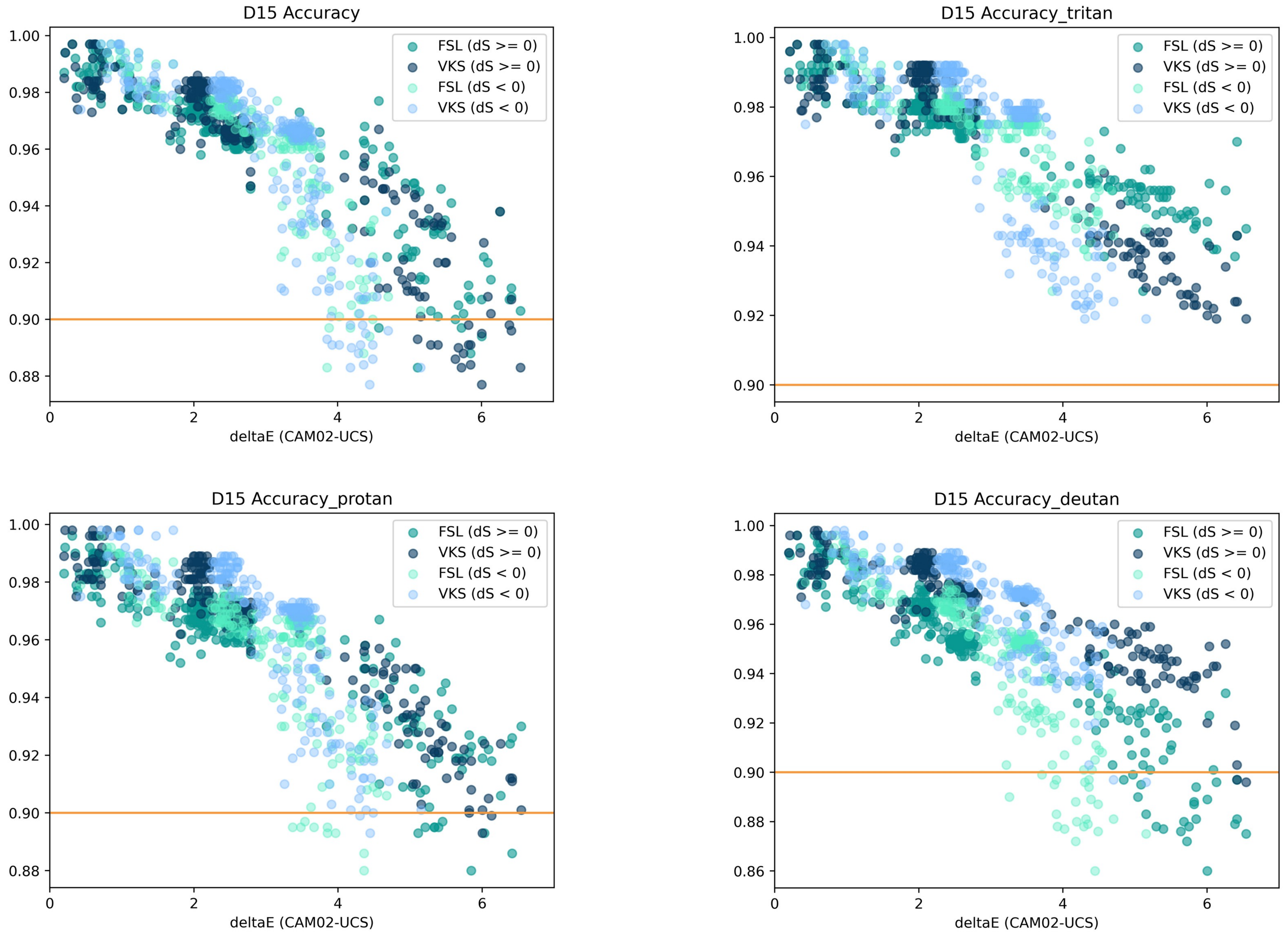
D15 Accuracy
D15 Accuracy_tritan
D15 Accuracy_protan
D15 Accuracy_deutan
FSL (dS >= 0)
VKS (dS >= 0)
FSL (dS < 0)
VKS (dS < 0)
deltaE (CAM02-UCS)
1.00
0.98
0.96
0.94
0.92
0.90
0.88
0.86
0
2
4
6

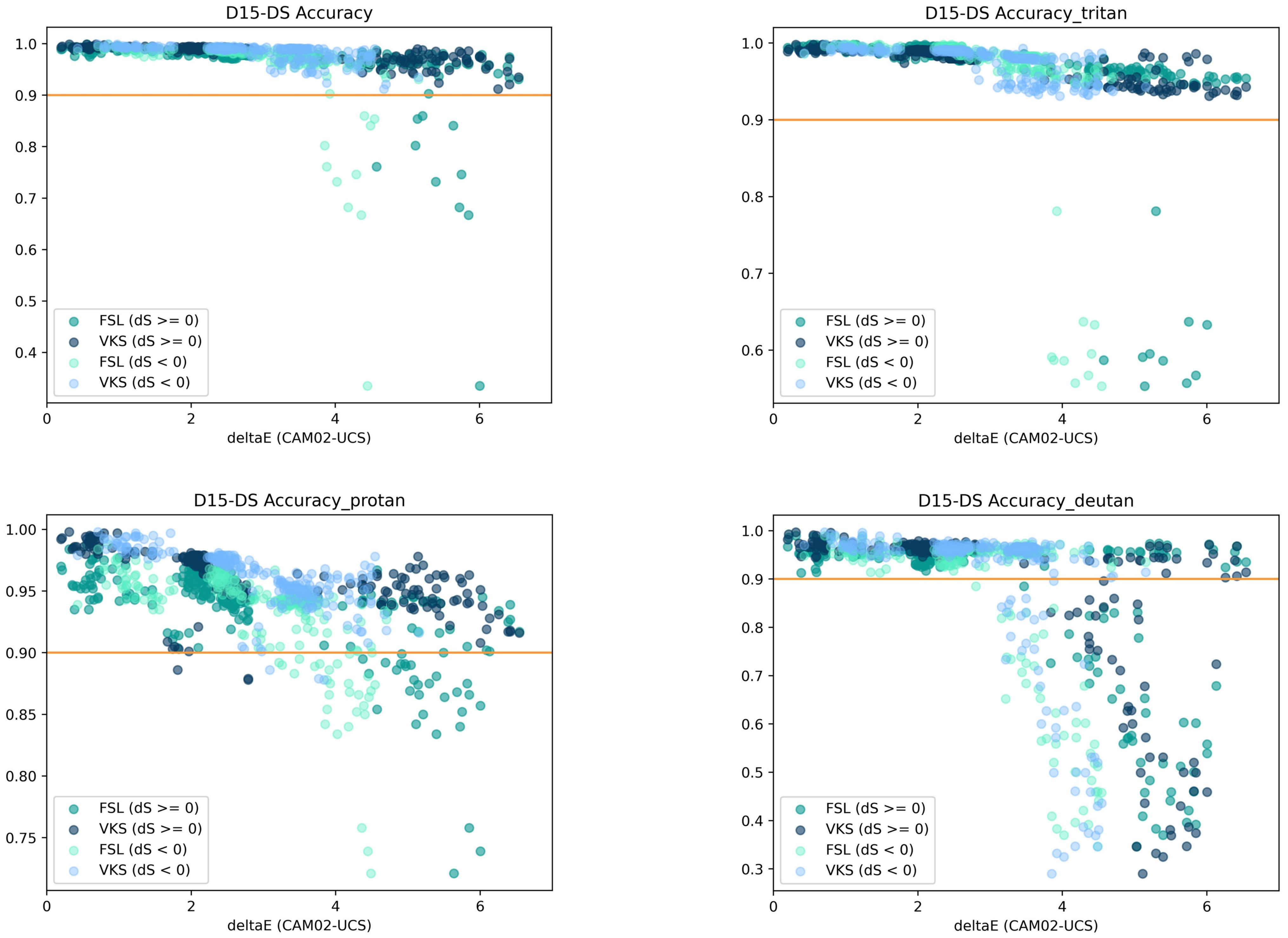

D15-DS Accuracy
D15-DS Accuracy_tritan
D15-DS Accuracy_protan
D15-DS Accuracy_deutan
FSL (dS >= 0)
VKS (dS >= 0)
FSL (dS < 0)
VKS (dS < 0)
deltaE (CAM02-UCS)

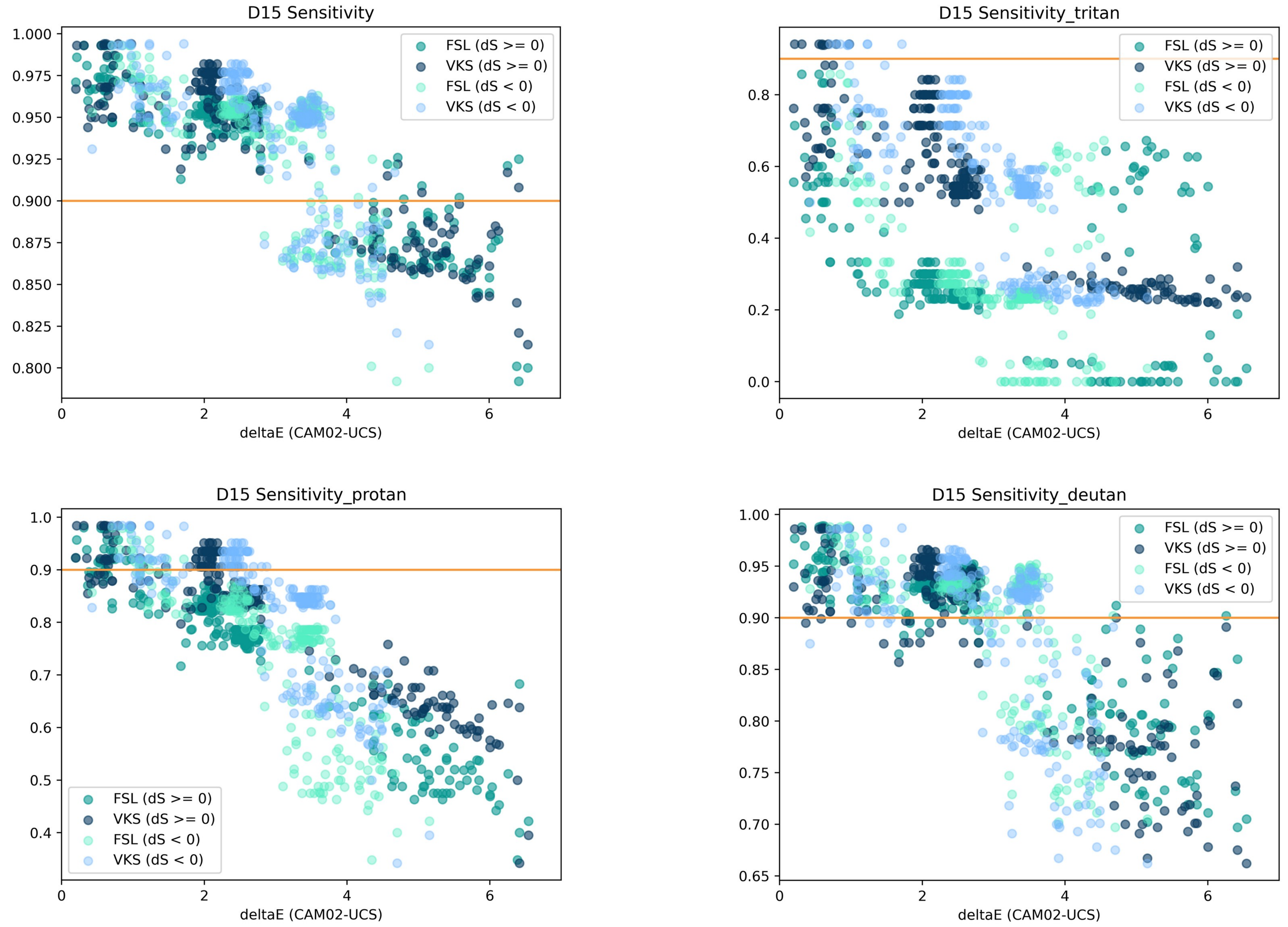
D15 Sensitivity
D15 Sensitivity_tritan
D15 Sensitivity_protan
D15 Sensitivity_deutan
FSL (dS >= 0)
VKS (dS >= 0)
FSL (dS < 0)
VKS (dS < 0)
deltaE (CAM02-UCS)

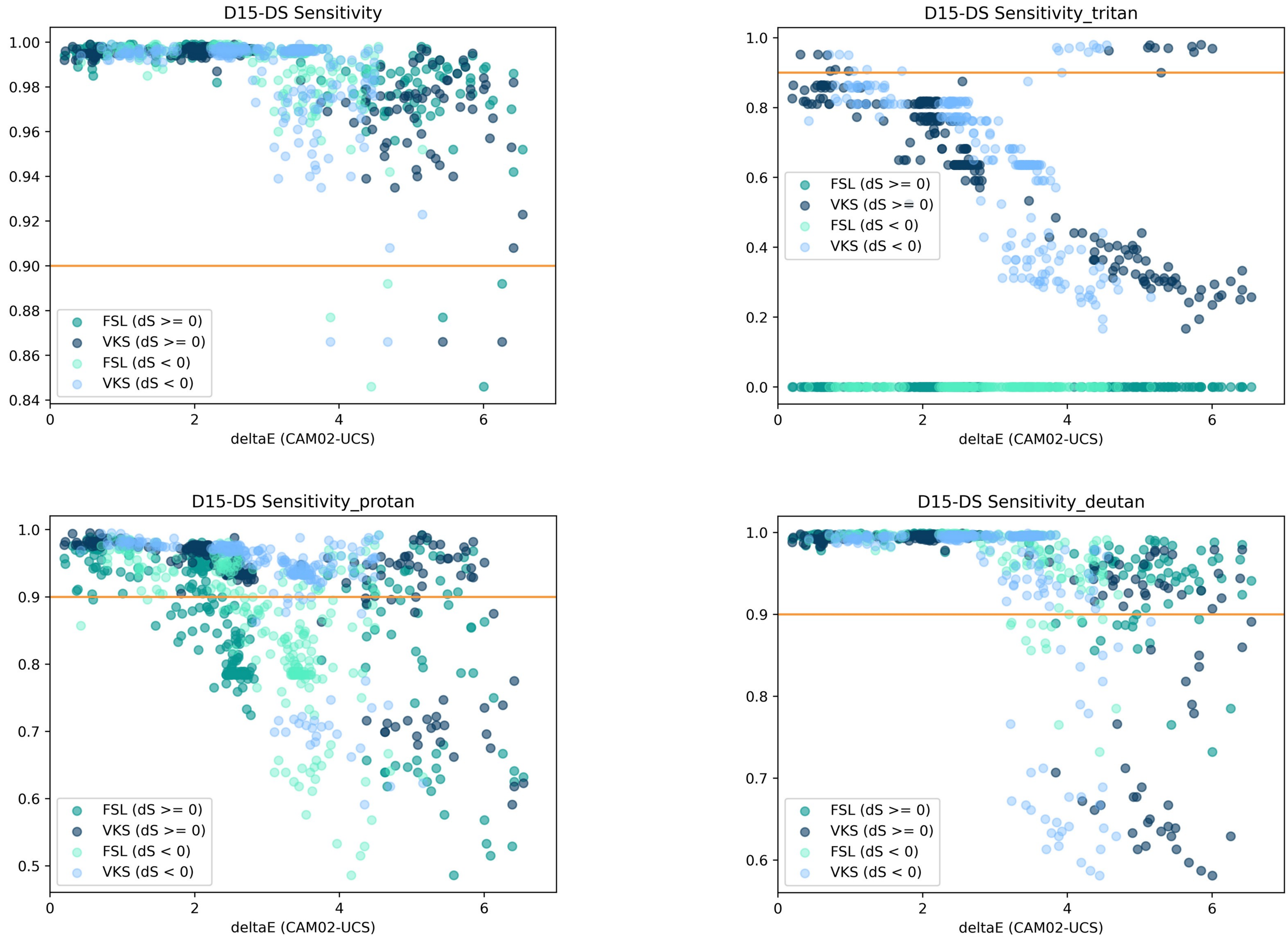
D15-DS Sensitivity
D15-DS Sensitivity_tritan
D15-DS Sensitivity_protan
D15-DS Sensitivity_deutan
FSL (dS >= 0)
VKS (dS >= 0)
FSL (dS < 0)
VKS (dS < 0)
deltaE (CAM02-UCS)
1.00
0.98
0.96
0.94
0.92
0.90
0.88
0.86
0.84
1.0
0.8
0.6
0.4
0.2
0.0
0.9
0.7
0.5
0
2
4
6

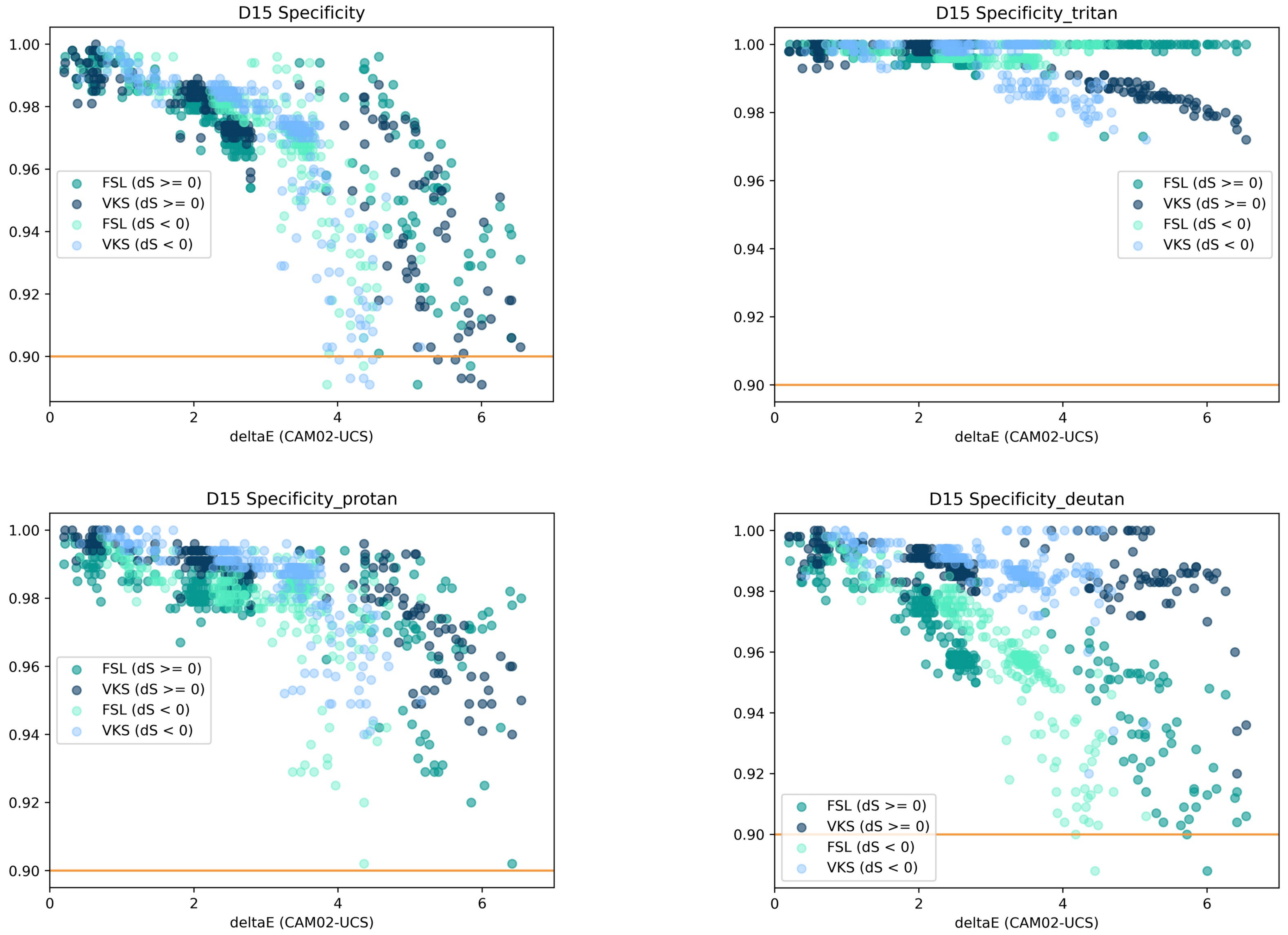
D15 Specificity
D15 Specificity_tritan
D15 Specificity_protan
D15 Specificity_deutan
FSL (dS >= 0)
VKS (dS >= 0)
FSL (dS < 0)
VKS (dS < 0)
deltaE (CAM02-UCS)
1.00
0.98
0.96
0.94
0.92
0.90
0
2
4
6

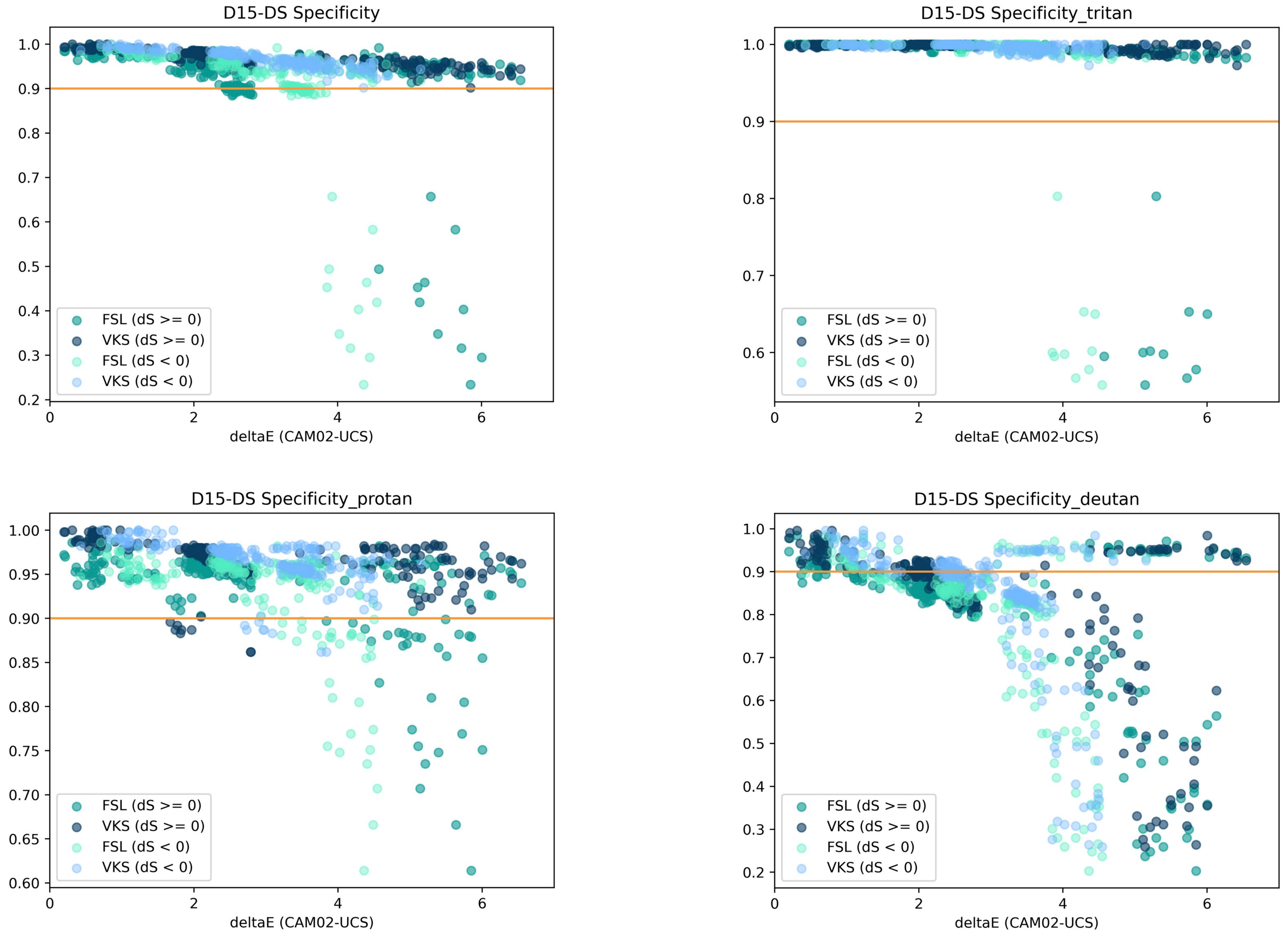
D15-DS Specificity
D15-DS Specificity_tritan
D15-DS Specificity_protan
D15-DS Specificity_deutan
FSL (dS >= 0)
VKS (dS >= 0)
FSL (dS < 0)
VKS (dS < 0)
deltaE (CAM02-UCS)

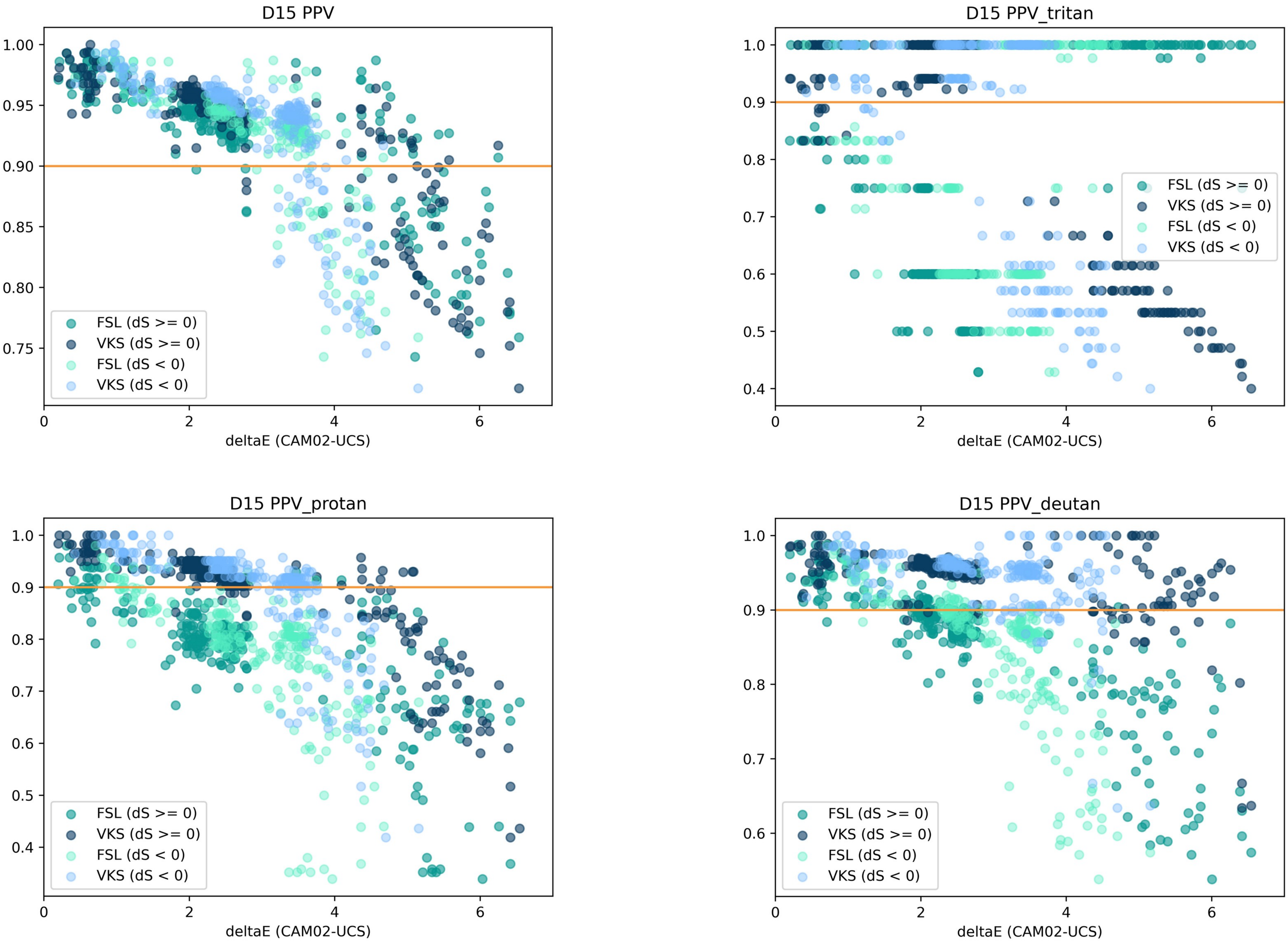
D15 PPV
D15 PPV_tritan
D15 PPV_protan
D15 PPV_deutan
deltaE (CAM02-UCS)
FSL (dS >= 0)
VKS (dS >= 0)
FSL (dS < 0)
VKS (dS < 0)

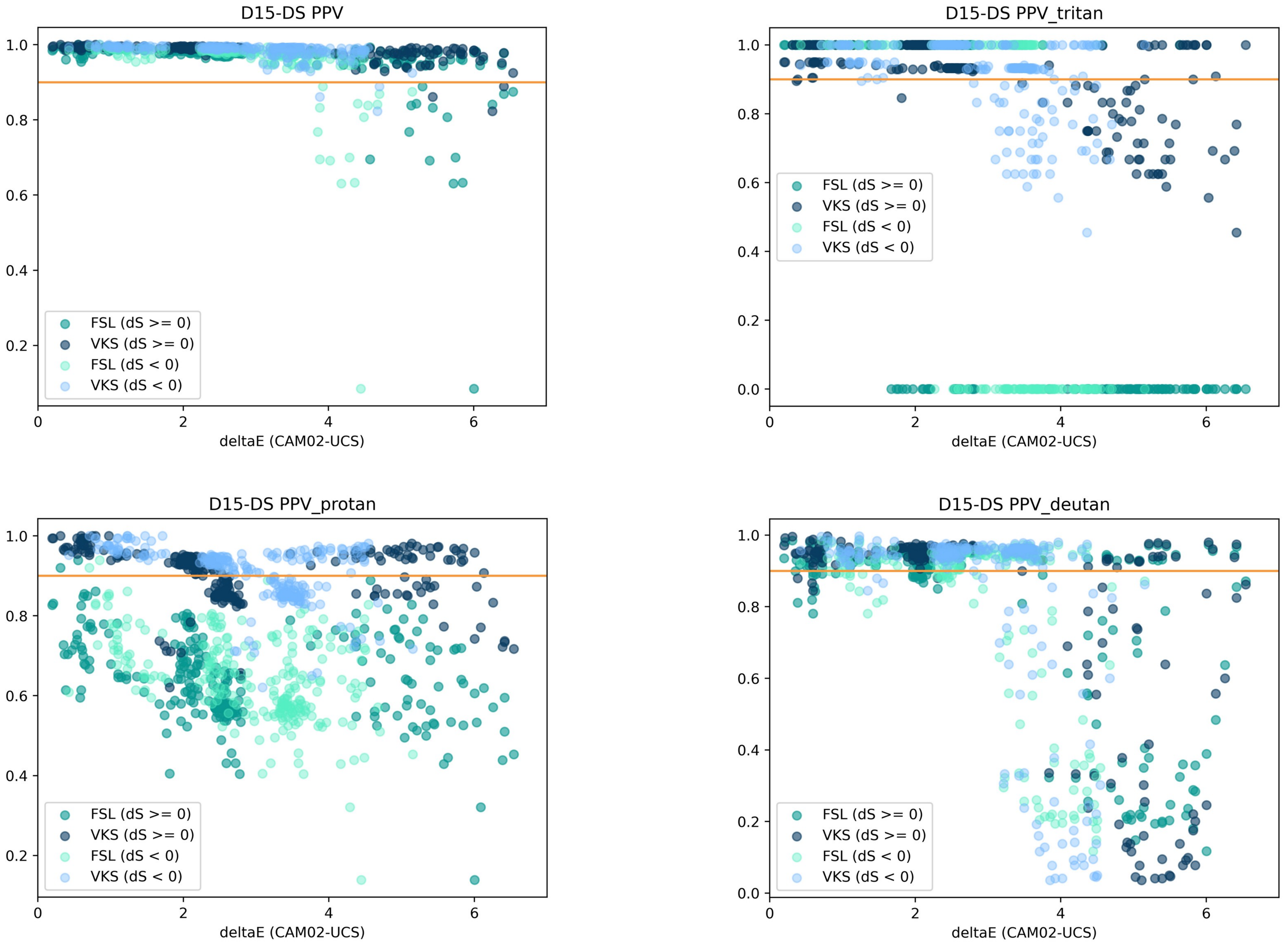

D15-DS PPV
D15-DS PPV_tritan
D15-DS PPV_protan
D15-DS PPV_deutan
FSL (dS >= 0)
VKS (dS >= 0)
FSL (dS < 0)
VKS (dS < 0)
deltaE (CAM02-UCS)

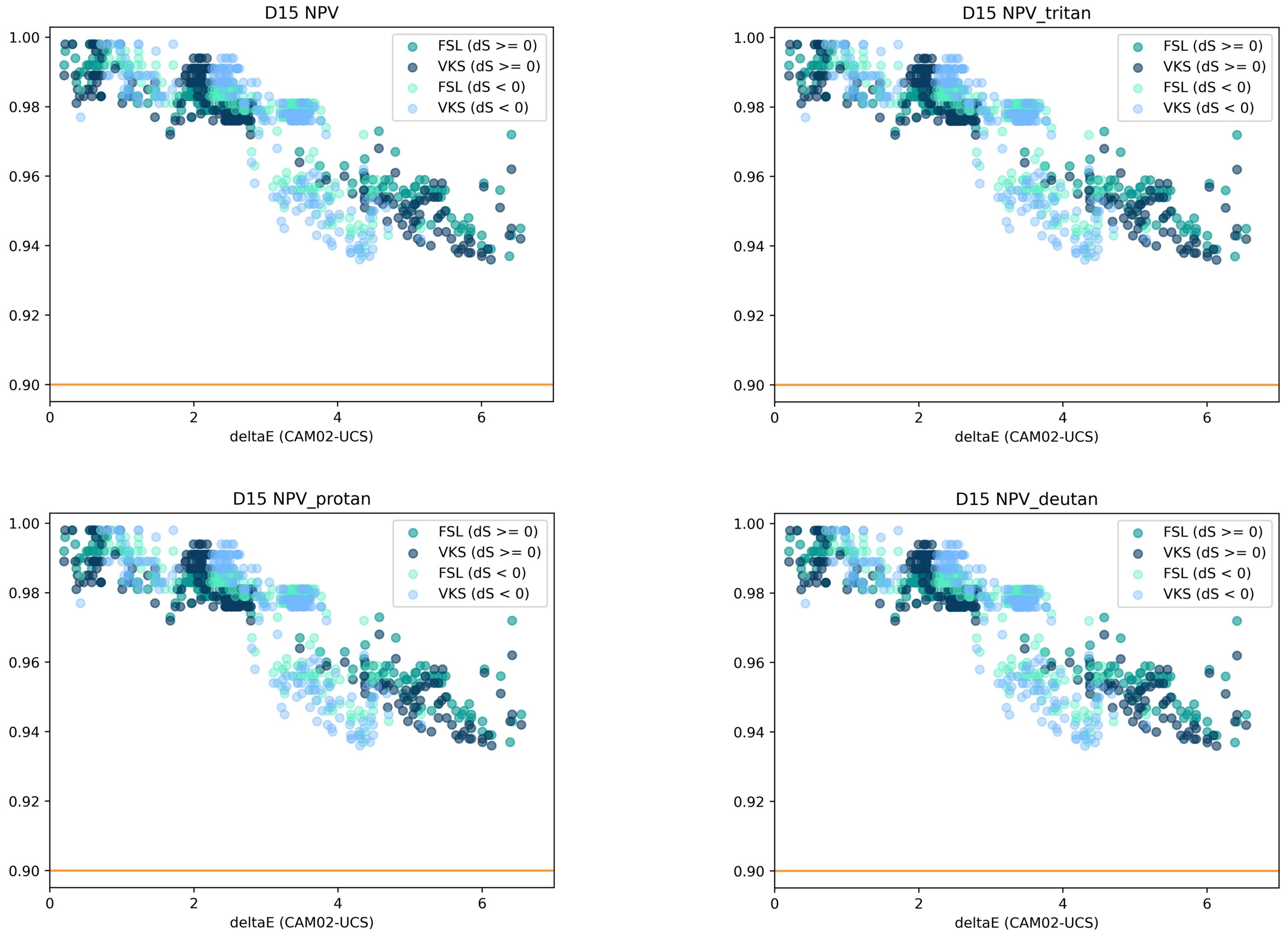
D15 NPV
D15 NPV_tritan
D15 NPV_protan
D15 NPV_deutan
FSL (dS >= 0)
VKS (dS >= 0)
FSL (dS < 0)
VKS (dS < 0)
1.00
0.98
0.96
0.94
0.92
0.90
0
2
4
6
deltaE (CAM02-UCS)

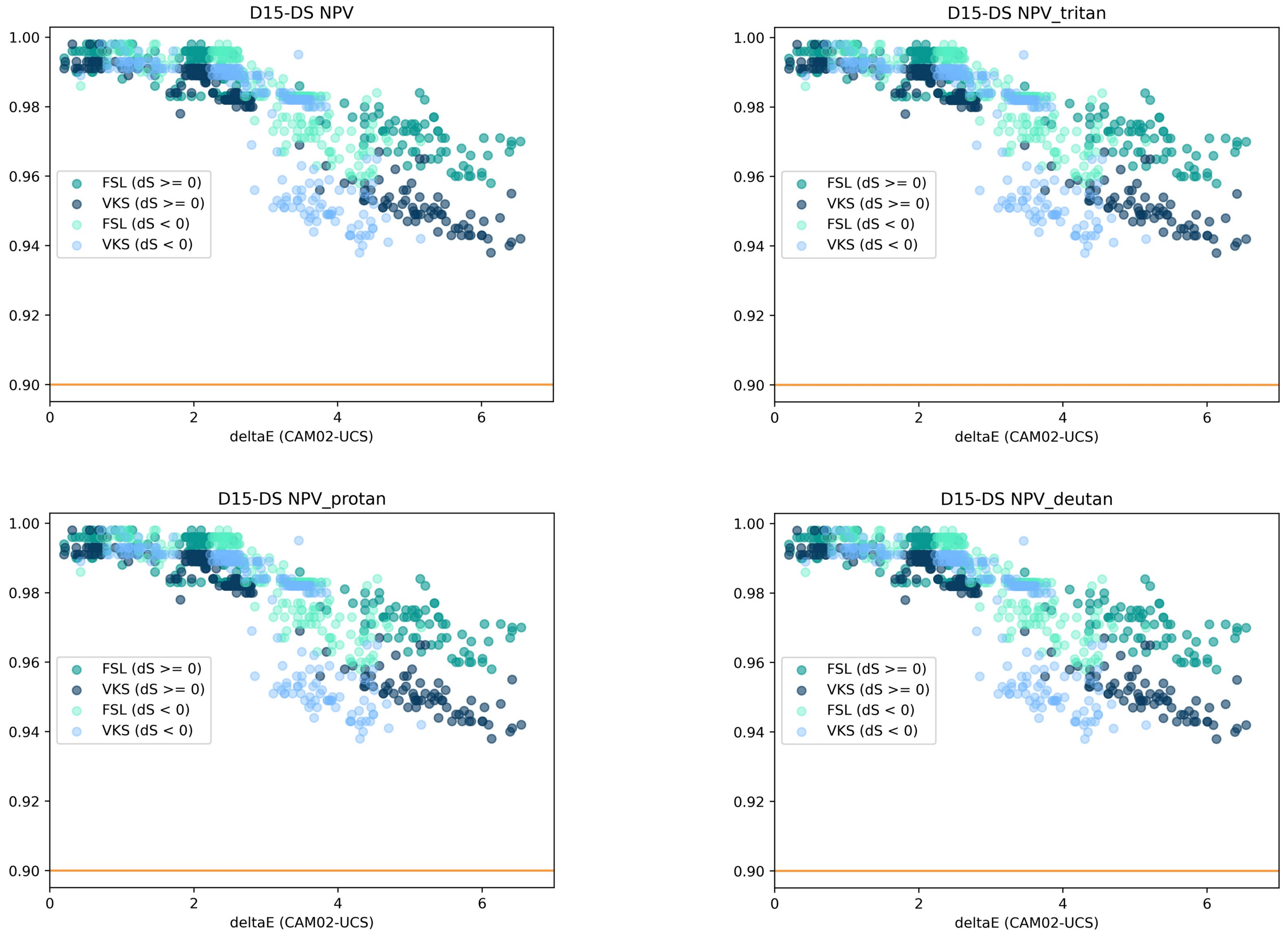
D15-DS NPV
D15-DS NPV_tritan
D15-DS NPV_protan
D15-DS NPV_deutan
FSL (dS >= 0)
VKS (dS >= 0)
FSL (dS < 0)
VKS (dS < 0)
deltaE (CAM02-UCS)
